# Welsh hospital admissions due to dog bites and strikes (2014-2022)

**DOI:** 10.1101/2024.03.05.24303782

**Authors:** Sara C Owczarczak-Garstecka, James A Oxley, John SP Tulloch

## Abstract

**Objective:** To describe the incidence and victim demographics of Welsh hospital admissions due to dog bites and strikes from 2014 to 2022.

**Study design:** Descriptive analysis of Welsh hospital admissions data.

**Methods:** Residents of Wales admitted to a Welsh National Health Service (NHS) hospital for a dog bite or strike, were identified using ICD-10 codes. The annual incidence of dog bite admissions between 2014 and 2022 was calculated and stratified by child–adult status, sex, and Local Health Boards. Trends over time were analysed using Chi-square test for trends.

**Results:** Hospital admissions due to dog bites and strikes have significantly increased from 16.3 per 100,000 to 23.7 per 100,000 population in 2022. This was driven by an increase in non-geriatric adults, and an increase in three Local Health Boards. The Swansea Bay area has the highest incidence in Wales (56.1 admissions per 100,000).

**Conclusions:** Wales has a higher incidence of dog bites than England or the Republic of Ireland. Work in the communities where incidence is exceptionally high is needed to understand the reasons behind having the highest incidence of dog bites in the British Isles and to establish the most efficacious methods for bite prevention.

## Introduction

Dog bites are a public health and epidemiological challenge due to the complexities of the human, dog and environmental interactions, the impact on the victims’ health and dog welfare.^1,2^ Despite no global estimate of dog bite incidence, the World Health Organisation (WHO) suggests that tens of millions of dog bite injuries occur annually, demonstrating the scale of the global health burden.^3^

In the United States of America (USA) non-fatal dog bites resulting in hospitalisations were estimated to cost at least $1.67 billion in 2021, 48% of this being medical costs, 9% work loss costs, and 43% quality of life loss costs.^4^ In England direct hospital healthcare costs were crudely estimated to be at least £70.8 million in 2017-2018,^5^ with costs to primary healthcare services, work loss costs, and quality of life costs remaining unknown. Additional societal costs, such as the work of the police and criminal justice system services have yet to be estimated.

Hospital admissions due to dog bites or strikes remain commonplace and have been increasing across the British Isles, with annual incidence increasing in both England and the Republic of Ireland (RoI). Dog bites follow different highly localised patterns,^5,6^ therefore public health resources need to be targeted at hotspot areas to maximise their cost-effectiveness. Within the United Kingdom (UK), this information is presently only available for England, as Northern Ireland, Scotland and Wales have not publicly published data on dog bite incidence trends, victim demographics or hotspots. Consequently, this study aims to describe the annual incidence of hospital admissions in Wales due to dog bites and strikes between 2014 and 2022, stratified by age, sex and region.

## Methods

Hospital admissions due to dog bites or strikes in Wales were explored utilising the admitted patient care (APC) datasets.^7^ The International Statistical Classification of Diseases and Related Health Problems 10th Revision (ICD-10) code W54 – ‘Bitten or struck by a dog’ was used to identify relevant summary data in the ‘external causes of injury – Welsh Residents’ tables.^8^ Data were extracted for eight financial years (April-March): 2014/2015 - 2021/2022.

The annual incidence of dog bite admissions in Wales was calculated and stratified by child– adult status and sex using the Office for National Statistics (ONS) mid-year population estimates as the denominator population.^9^ The annual incidence was stratified by child– adult status and by sex. Due to the age-bands in the APC data, a child was defined as anyone age ≤14 years of age. Individuals aged 15-18 years of age could not be defined as children as they were included in the 15-59 age band. Cases age 60-74 were classed as older adults and cases over the age of 75 were categorised as geriatric adults. The annual incidence in each Local Health Board (LHB) was calculated and mapped; this was based on the patient’s residence rather than geographic location where they were bitten. LHBs are responsible for the planning and delivery of National Health Services in each respective area.^10^ Trends in incidence were explored using a Chi-squared test for trend. A summary of management data, such as mean and median length of stay, and number of bed days were also described.

All statistical and spatial analyses were carried out using R language (version 3.2.0; R Core Team 2015). The results were deemed statistically significant where p<0.05. No ethical approval was needed, as this was an analysis of publicly available data, with no personally identifiable information. Patients were not involved in the design, nor participated in the research, of this study.

## Results

Between the financial years 2014/2015 and 2021/2022, there were 4,754 finished consultant episodes with the ‘bitten or struck by a dog’ ICD-10 code. There was a total of 7,536 bed days, with a median length of stay of one day, and the overall mean length of stay of 1.6 days.

There was a significant increase in dog bite hospital admissions (12 = 48.1, p<0.001) from 15.3 (95%CI 14.0-16.8) admissions per 100,000 population in 2014/2015 to a peak of 23.7 (95%CI 22.0-25.4) admissions per 100,000 in 2021/2022 (Fig 1).

**Figure 1.**
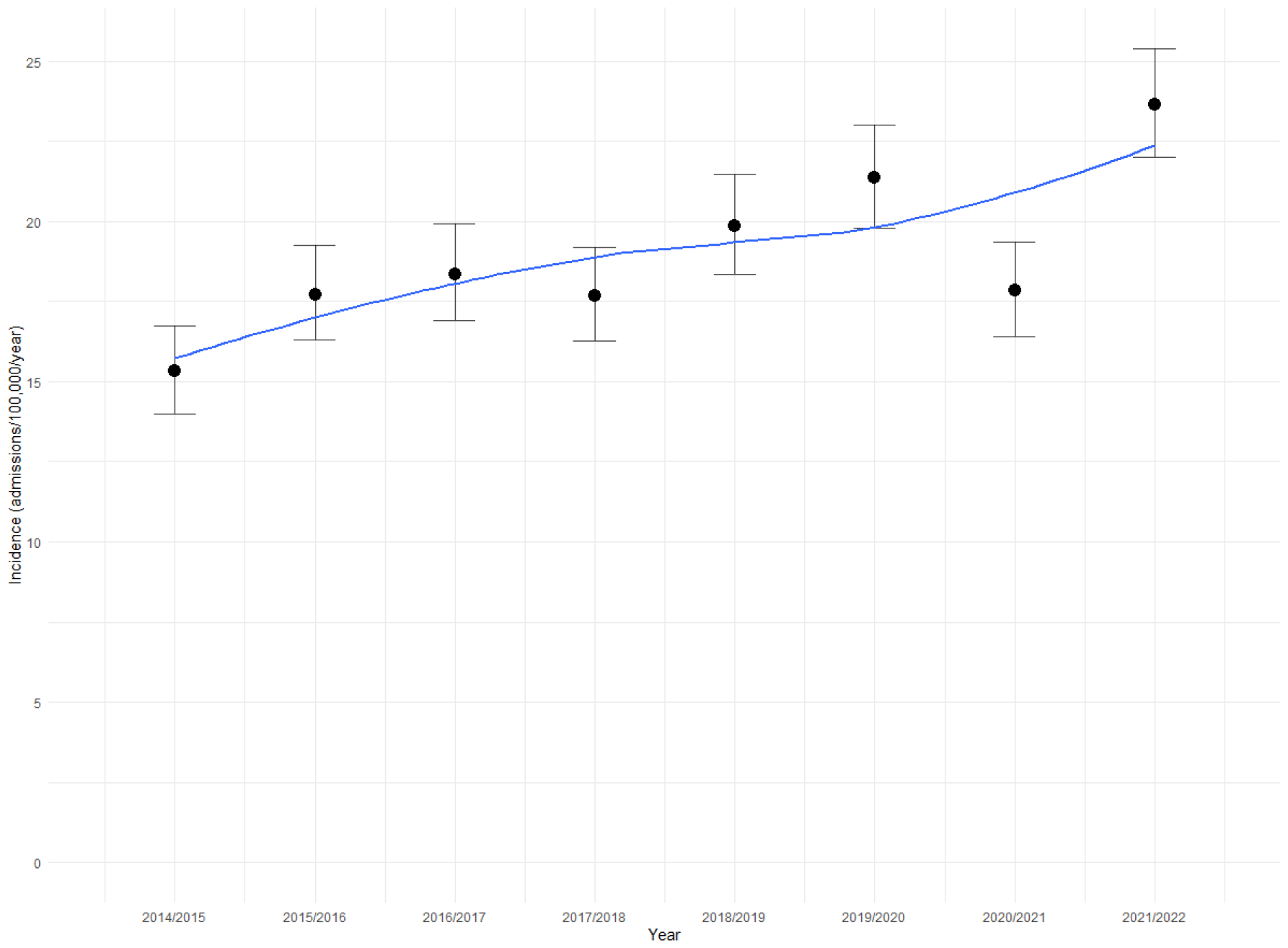
Annual incidence of hospital admissions due to ‘dog bites and strikes’ in Wales.

Children (≤14 years) made up 20.8% (95%CI 19.7-22.0) of all dog bite hospital admissions. The mean annual incidence for children was 23.6 (95%CI 22.2-25.2) admissions per 100,000 population, with a minimum incidence of 20.6 (95%CI 17.0-24.8) in 2015/2016 and a maximum of 27.5 (95%CI 23.3-32.2) in 2018/2019. The incidence of dog bite admissions for children showed no significant increase or decrease (12 = 0.71, p = 0.40) within the study period (Fig 2).

**Figure 2.**
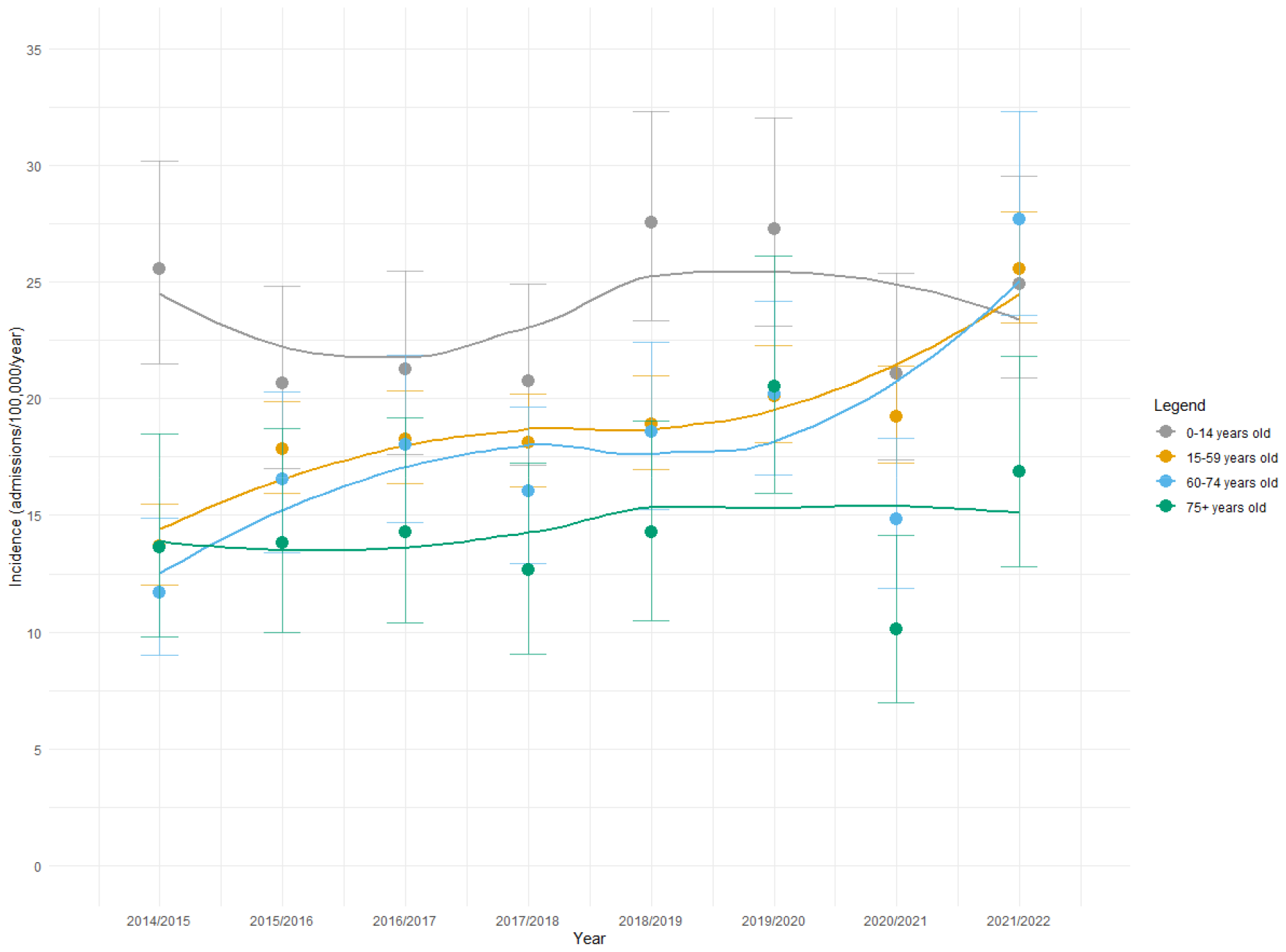
Annual incidence of hospital admissions due to ‘dog bites and strikes’ in Wales, stratified by age group.

In contrast, there was a significant increase of dog bite admissions in adults (□2 = 69.6, p<0.001) from 13.3 (95%CI 11.9-14.7) in 2014/15 to 24.9 (95%CI 23.1-26.9) in 2021/2021. When stratified by age band the increase was not significant in 75-year-olds and over (⍰2 = 0.74, p = 0.39). A significant increase was observed in 15–59-year-olds (⍰2 = 51.0, p<0.001) and 60-74 year-olds (⍰2 = 22.9, p<0.001). Over half of all adult admissions were female, 52.5% (95%CI 51.1-54.0), this proportion was stable over the study period. Men showed a significant increase in incidence (⍰2 = 13.2, p<0.001) from 15.6 (95%CI 13.8-17.8) cases per 100,000 in 2014-2015 to 21.8 (95%CI 19.6-24.3) in 2021-2022. Likewise, women showed a significant increase in incidence (⍰2 = 35.0, p<0.001) from 15.1 (95%CI 13.3-17.1 cases per 100,000) in 2014/2015 to 25.9 (95%CI 23.5-28.5) in 2021/2022.

Almost all records had information about which LHB the patient was a resident in (99.4%, n = 4726). Due to changes in health board boundaries, trends could only be assessed in five health boards. Of these, only Betsi Cadwaladr University Health Board (UHB) (the most northern region) had a significant change in incidence, rising from 10.9 in 2016/2017 to 19.1 in 2020/2021 (⍰2=14.0, p<0.001) (Fig 3).

**Figure 3.**
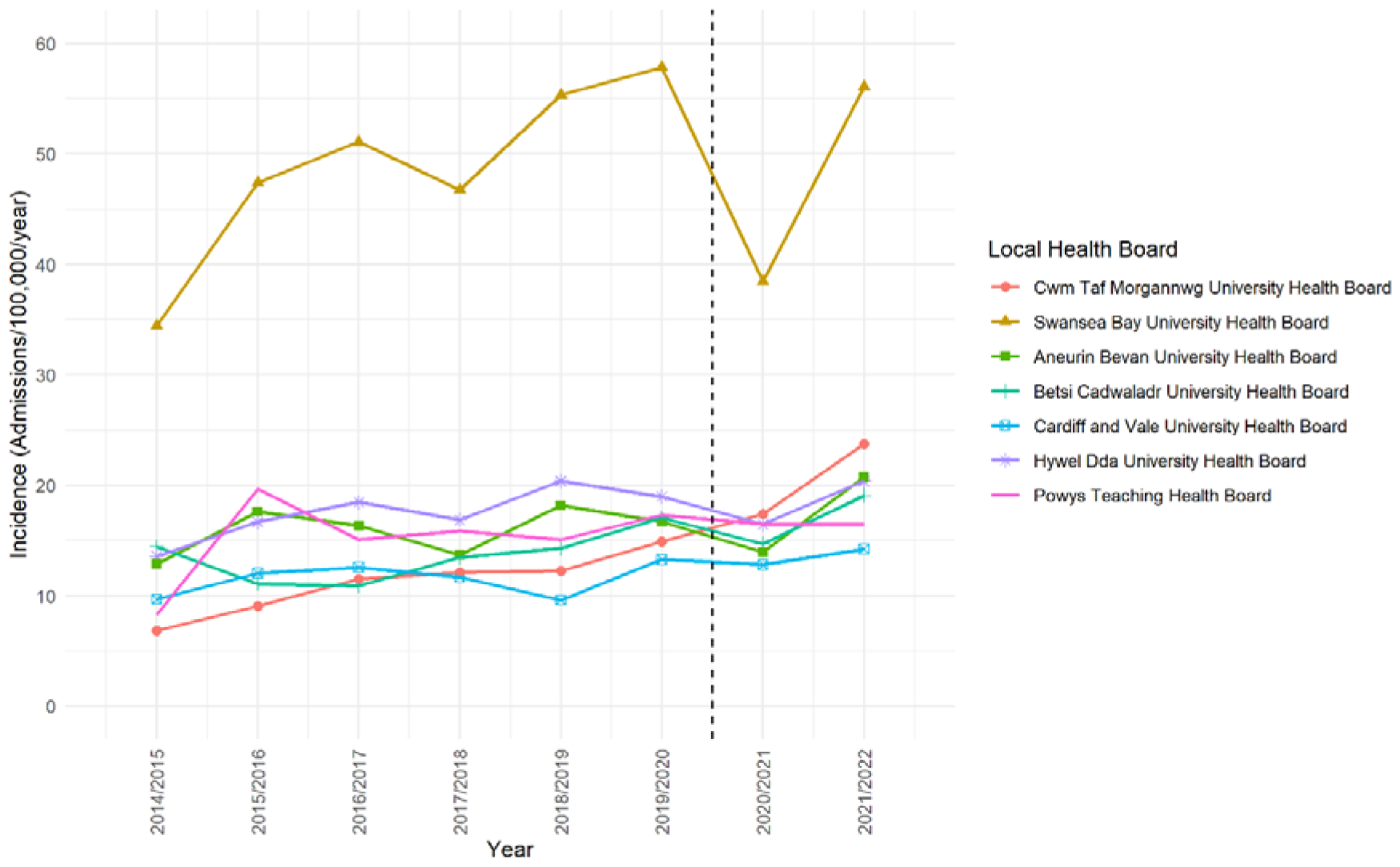
Annual incidence of hospital admissions due to ‘dog bites and strikes’ in Wales, stratified by Local Health Board of patients’ residence. (Dashed line represents changes in LHB boundaries)

After 2019 LHB boundary changes, Bridgend local authority moved from Abertawe Bro Morgannwg UHB to Cwm Taf UHB. Abertawe Bro Morgannwg UHB was renamed Swansea Bay UHB, and Cwm Taf UHB was renamed Cwm Taf Morgannwg UHB. Prior to these changes the region with the highest incidence was Abertawe Bro Morgannwg UHB in 2018/2019 (55.3, 95%CI 48.4-63.2), after 2019, the resultant smaller region of Swansea Bay UHB had the highest incidence of 57.8 (95%CI 50.8-65.9) admissions per 100,000 in 2019/2020. If Abertawe Bro and Swansea Bay were viewed as the same area the increase would have reached statistical significance (⍰2=8.7, p<0.01), likewise with Cwm Taf (⍰2=56.6, p<0.001). Incidence of dog bite admissions by LHB for the year 2021/2022 are shown in Figure 4.

**Figure 4.**
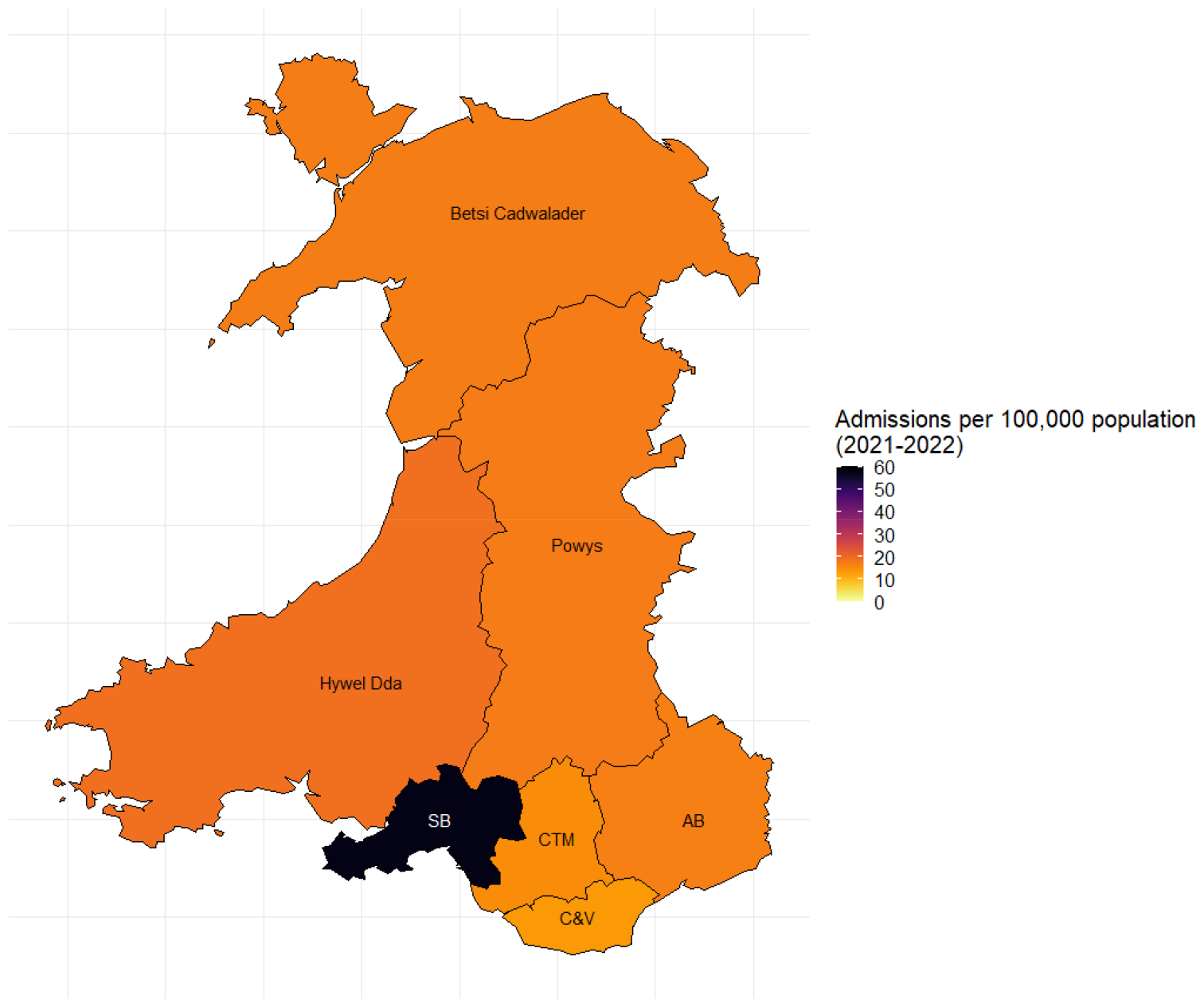
The 2021-2022 incidence of hospital admission due to ‘dog bites and strikes’ in Wales, stratified by Local Health Board of patients’ residence (SB= Swansea Bay, CTM= Cwm Taf Morgannwg, AB= Abertawe Bro Morgannwg, C & V= Cardiff and Vale).

## Discussion

This study highlights that Wales has a growing dog bite problem. Between 2014/2015 and 2021/2022, the incidence of Welsh dog bite hospital admissions increased by 54.9%. This is greater than the growth in incidence seen in RoI (50% increase) and England (14.7% increase) over the same time period.^11,12^ The increase appears to be driven by a raise in bites to non-geriatric adults, and in three specific regions of the country. The Welsh national incidence (24.9 cases per 100,000 in 2021/2022) is higher than the incidence in England (14.8 cases per 100,000 in 2021/22) or Ireland (8.7 cases per 100,000 in 2021/22).

Multiple factors can be postulated as drivers of the rise in dog bite incidence. The sourcing of dogs has changed over the last decade. Although exact estimates are not available, a substantial proportion of the UK pet-dog population is believed to have been smuggled into the UK from low-quality breeding farms (“puppy farms”) abroad or bred in the UK in sub-optimal welfare conditions.^13,14^ In the UK, Wales has the highest density of puppy farms per capital.^15^ According to a survey of 745 veterinary professionals, the lack of research prior to dog acquisition is regarded as one of the top five dog welfare concerns.^16^ In fact just 54% of 8381 current UK dog owners carried out any pre-acquisition research,^17^ and in a separate study of 142 owners half of dog acquisitions were unplanned, i.e. spontaneous.^18^ These factors are important as lack of enriched and safe early-life environment and adequate, structured socialisation of puppies is a risk-factors for multiple behaviour and health problems, including human-directed aggression.^19–22^ It has been hypothesised that the rise in the pet-dog population post the COVID-19 pandemic (an estimated 2.5 million dogs were acquired during the COVID-19 pandemic,^23^ the pre-pandemic population was estimated at 12.6 million dogs in 2019 ^24^) has led to a rise in dog bites, due to the increasing volume of human-dog interactions. However, it must be noted that there is no discernible change in the trajectory of dog bite incidence post COVID-19. Not meeting a dog’s exercise needs and thus not providing them with adequate socialisation opportunities has also been associated with higher risk canine anxiety, which in turn is a risk factor for human-directed aggression.^25^ Finally, it has been hypothesised that depiction of dogs in popular and social media contributes to the popularisation of an anthropomorphic view of dogs.^26^ This view has been associated with changes in expectations of dog behaviour and engagement in behaviours that carry a high risk of being bitten (e.g. expecting a dog to enjoy being hugged, which some dogs may find intimidating).^27,28^

The incidence rate of dog bite-related hospitalisation has nearly doubled within the Welsh adult population, excluding geriatric adults. Alongside England and RoI, Wales is now the third nation where the national population increase in dog bites is being driven by adult cases, making this an emerging trend. Similar to England,^5^ the incidence in adults is now on par with children, who are traditionally viewed as the demographic with the highest incidence of dog bites.^2^ In Wales, the incidence in children remain stable but high, at a level that it is almost double of the incidence seen in children in England.^5^ While the incidence of all accidental injuries to children remains stable across the UK nations since 2013, the Welsh rates remain higher than rates in England or Scotland (due to differences in data, comparisons with Northern Ireland are not possible)^29^, thus higher rates of bites may be a part of a broader pattern. In all nations where the raise in dog bites has been driven by dog bites to adults, bite prevention initiatives primarily target children; possibly contributing to the observed changes.^30,31^

Similar to RoI,^12^ but by contrast to England where the incidence of bites is higher in men,^5^ in Wales there were no differences in incidence of dog bites stratified by sex. Conclusions cannot easily be drawn about this without the data being analysed in a disaggregated manner.

Most areas of Wales are not seeing an increase in dog bites, but their base line levels do appear to be higher than the majority of local authorities in England (mean of 8.0 dog bite admissions per 100,000 population per year). There are three areas that are increasing: Swansea Bay UHB, Cwm Taf UHB, and Betsi Cadwaladr UHB. Cym Taf and Betsi Cadwalader both have an incidence which is equivalent to being in the top 10 local authorities of England, which ranges from 16.0-24.2 dog bite admissions per 100,000 population per year. Swansea Bay has an incidence that is more than two times higher than anywhere else in England or Wales. The top local authorities in England tend to be areas with high levels of socio-economic deprivation, with predominately white populations.^5^ In Scotland, dog bites have also been seen in increasing incidence in areas with higher levels of socio-economic deprivation.^6^ Swansea Bay is predominately white (96.1%)^32^ and is an area with some of the highest levels of deprivation in Wales and in the UK,^33^ with 38% of locations deemed to have deep-rooted deprivation.^34^ It is possible that Swansea Bay shares similar characteristics as other areas with high levels of dog bites. However, these are currently assumptions and without disaggregated data analysis this cannot be proven.

It is also unclear why dog bites are higher in areas of higher socio-economic deprivations. In these areas, dog owners may be less able to cover the costs of veterinary and behavioural support. Untreated pain is a risk factor for human-directed aggression,^35,36^ and dogs bred on puppy farms are more likely to suffer with poor health.^37,38^ It is likely that in more deprived areas properties are smaller resulting in some dogs experiencing emotional conflict as they cannot avoid unwanted interactions due to the confined space. It is also possible that dogs are more likely to be acquired from puppy farms or “backstreet breeders” due to lower costs. As explained earlier, source of puppy acquisition and puppy’s early life environment are among the main risk factors for human-directed aggression. Backstreet breeding has been linked to the European illegal puppy trade, which has strong links to organised crime and breeding dogs for fighting, intimidation and/or personal protection. ^39–43^ Presence of dogs trained to attack or encouraged to react aggressively to unfamiliar people (commonly referred to as status dogs) can contribute to bites to unintended victims in the surrounding communities.^41,43^ However, the scale of using dogs in criminal activities and its impact on communities is unknown. Without further targeted local investigations, reasons for bites in socio-economically deprived localities in Wales will remain unknown.

Assuming that the minimum healthcare cost of a dog bite admission is £406 (the cost of “regular admission”), then dog bites have cost the NHS in Wales at least £1.93 million over the study period, and over £300,000 in the 2021/2022.^44^ The true cost is likely to be significantly higher, the Centre for the Disease Control and Prevention in the USA estimates that in 2021 the average medical cost of a non-fatal hospital admission for a dog bite was $54,196.^45^ It is possible that dog bites are costing the NHS in Wales millions of pounds annually, without accounting for the quality-of-life costs to those who have been injured, or additional societal costs of dog bites outlined earlier.

The major limitation of this study is that it is an analysis of summary data rather than disaggregated raw data, as such we cannot model different demographic variables to understand their relative association with the increasing incidence. The ICD-10 codes used to identify the dog bites and strikes were only available at the three-character category level, rather than the four-character subcategory level, and so we could not identify the location of the injury (i.e. home, street) that would be provided with this subcategory.^5,8^ Previous literature has highlighted the issue that this code combines two very distinct injury mechanisms (bite or strike), and without details of the resultant trauma it has difficult to ascertain the relative prevalence of each. However, it appears that bites tend to be more prevalent than strikes.^5^ Due to the nature of this aggregated data the geographic resolution is not high, and we cannot understand the impact of socio-economic deprivation nor rurality on dog bites. As with all hospital records research, the years 2020 and 2021 hospital services were severely affected by the COVID-19 pandemic, and thus data for the financial year 2020-2021 may not be reflective of a standard year.^46–48^ Similar to other research on electronic health records, no data is provided about the context of the events surrounding the injury, nor any information about the dog involved. Understanding of these factors is critical to develop effective dog bite prevention strategies.

## Conclusions

Dog bites in Wales are increasing at a higher and faster rate than in England or Ireland. This has been driven by an increase in dog bites to non-geriatric adults. Concerningly Wales may contain a region with the highest dog bite incidence in the United Kingdom; Swansea Bay. This highlights the need for further investigation to understand why dog bites are so prevalent here. Any dog bite prevention or intervention strategies need to be targeted to the worst-affected areas to be resource efficient and to have the greatest impact on reducing dog bites and thus improving the health of the public.

## Data Availability

All data produced in the present work are contained in the manuscript. The raw open data is available at https://dhcw.nhs.wales/information-services/information-delivery/hospital-admissions/hospital-admissions-annual-online-data-tables-statistical-reports/

## Statement of ethical approval

No ethical approval was needed as this was an analysis of publicly available data, with no personally identifiable information. Patients were not involved in the design, nor participated in the research, of this study.

## References

1. Colmers-Gray IN, Tulloch JS, Dostaler G, Bai AD. Management of mammalian bites. BMJ. 2023 Feb 2;e071921. 10.1136/bmj-2022-071921

2. Jakeman M, Oxley JA, Owczarczak-Garstecka SC, Westgarth C. Pet dog bites in children: Management and prevention. BMJ Paediatr Open. 2020;4(1):15–8. 10.1136/bmjpo-2020-000726

3. World Health Organization. Animal bites. https://www.who.int/news-room/fact-sheets/detail/animal-bites; 2024 [accessed 1 February 2024]

4. CDC. Injury Prevention & Control - Data and Statistics (WISQARS). https://www.cdc.gov/injury/wisqars/index.html; 2024 [accessed 1 February 2024]

5. Tulloch JSP, Owczarczak-Garstecka SC, Fleming KM, Vivancos R, Westgarth C. English hospital episode data analysis (1998 – 2018) reveal that the rise in dog bite hospital admissions is driven by adult cases. Sci Rep. 2021;11(1):1767. 10.1038/s41598-021-81527-7

6. Hooper J, Lambert P, Buchanan-Smith H, Robertson T. Exploring Social and Locality Variations of Dog Bites in Scotland Using Administrative Data Sources. Int J Popul Data Sci. 2022 Aug 25;7(3):1810. 10.23889/ijpds.v7i3.1810

7. Digital Health and Care Wales. Hospital Admissions: Annual Online Data Tables - Statistical Reports. https://dhcw.nhs.wales/information-services/information-delivery/hospital-admissions/hospital-admissions-annual-online-data-tables-statistical-reports/; 2024 [accessed 1 February 2024]

8. World Health Organisation. ICD-10 Version:2019 https://icd.who.int/browse10/2019/en; 2020 [accessed 1 February 2024]

9. Office for National Statistics. Population estimates for England and Wales: mid-2022. https://www.ons.gov.uk/peoplepopulationandcommunity/populationandmigration/populationestimates/bulletins/populationestimatesforenglandandwales/mid2022; 2023 [accessed 1 February 2024]

10. NHS Wales. Local Services. https://www.nhs.wales/hpb/local-services/; 2023 [accessed 1 February 2024]

11. NHS England. Hospital admissions caused by dog bites. https://digital.nhs.uk/supplementary-information/2023/hospital-admissions-caused-by-dog-bites; 2023 [accessed 1 February 2024]

12. Ryan E, O’Farrell A, O’Suillleabhain--McKeown D. Trends in in-hospital admissions due to dog bites in Ireland from 2012–2021. Ir Med J. 2023;116(10):P887.

13. Dogs Trust. Puppy Smuggling. https://www.dogstrust.org.uk/downloads/2020_Puppy_smuggling_report.pdf; 2020 [accessed 1 February 2024]

14. Wyatt T, Maher J, Biddle P. Scoping Research on the Sourcing of Pet Dogs From Illegal Importation and Puppy Farms 2016-17. https://www.gov.scot/publications/scoping-research-sourcing-pet-dogs-illegal-importation-puppy-farms-2016/documents/; 2017 [accessed 1 February 2024]

15. Ross KE, Langford F, Pearce D, McMillan KM. What Patterns in Online Classified Puppy Advertisements Can Tell Us about the Current UK Puppy Trade. Animals. 2023 May 18;13(10):1682. 10.3390/ani13101682

16. PDSA. PDSA Animal Wellbeing Report 2023. https://www.pdsa.org.uk/what-we-do/pdsa-animal-wellbeing-report/paw-report-2023; 2023 [accessed 1 February 2024]

17. Holland KE, Mead R, Casey RA, Upjohn MM, Christley RM. “Don’t Bring Me a Dog…I’ll Just Keep It”: Understanding Unplanned Dog Acquisitions Amongst a Sample of Dog Owners Attending Canine Health and Welfare Community Events in the United Kingdom. Animals. 2021 Feb 25;11(3):605. 10.3390/ani11030605

18. Mead R, Holland KE, Casey RA, Upjohn MM, Christley RM. “Do Your Homework as Your Heart Takes over When You Go Looking”: Factors Associated with Pre-Acquisition Information-Seeking among Prospective UK Dog Owners. Animals. 2023 Mar 10;13(6):1015. 10.3390/ani13061015

19. Gazzano A, Mariti C, Notari L, Sighieri C, McBride EA. Effects of early gentling and early environment on emotional development of puppies. Appl Anim Behav Sci. 2008 Apr;110(3–4):294–304. 10.1016/j.applanim.2007.05.007

20. Puurunen J, Hakanen E, Salonen MK, Mikkola S, Sulkama S, Araujo C, et al. Inadequate socialisation, inactivity, and urban living environment are associated with social fearfulness in pet dogs. Sci Rep. 2020 Feb 26;10(1):3527. 10.1038/s41598-020-60546-w

21. Mikkola S, Salonen M, Puurunen J, Hakanen E. Aggressive behaviour is affected by demographic, environmental and behavioural factors in purebred dogs. Sci Rep. 2021;(0123456789):1–10. 10.1038/s41598-021-88793-5

22. McEvoy V, Espinosa U, Crump A, Arnott G. Canine Socialisation: A Narrative Systematic Review. Animals. 2022 Oct 22;12(21):2895. Available from: 10.3390/ani13010081.

23. PDSA. PDSA Animal Wellbeing (PAW) Report. https://www.pdsa.org.uk/get-involved/our-campaigns/pdsa-animal-wellbeing-report. 2022 [accessed 1 February 2024]

24. McMillan KM, Harrison X, Upjohn MM, Christley RM, Casey RA. Estimation of the size, density, and demographic distribution of the UK pet dog population. Res Sq. 2024; 10.21203/rs.3.rs-3772889/v1

25. Tiira K, Lohi H. Early Life Experiences and Exercise Associate with Canine Anxieties. PLoS One. 2015 Nov 3;10(11):e0141907. 10.1371/journal.pone.0141907

26. Mota-Rojas D, Mariti C, Zdeinert A, Riggio G, Mora-Medina P, del Mar Reyes A, et al. Anthropomorphism and Its Adverse Effects on the Distress and Welfare of Companion Animals. Animals. 2021 Nov 15;11(11):3263. 10.3390/ani11113263

27. Meints K, Brelsford V, De Keuster T. Teaching Children and Parents to Understand Dog Signaling. Front Vet Sci. 2018 Nov 20;5(11):3263. 10.3389/fvets.2018.00257

28. Baatz A, Bidgood A, Taylor G, Young R. The trouble with a cuddle: Families’ experiences of supervising interactions between children in middle childhood and the family dog. Human-Animal Interact. 2023 Dec 7; 10.1079/hai.2023.0042

29. RCPCH. Accidental Injury. https://stateofchildhealth.rcpch.ac.uk/evidence/injury-prevention/accidental-injury/; 2021 [accessed 1 February 2024]

30. Dogs Trust. Dogs and children: living safely together. https://www.dogstrust.org.uk/dog-advice/life-with-your-dog/at-home/dog-and-child-safety; 2023 [accessed 1 February 2024]

31. Dogs Trust. Be Dog Smart! https://www.learnwithdogstrust.ie/be-dog-smart/; 2023 [accessed 1 February 2024]

32. StatsWales. Population estimates by local authority and ethnicity. https://statswales.gov.wales/Catalogue/Population-and-Migration/Population/Estimates/Ethnicity/PopulationEstimates-by-Localauthority-Ethnicity; 2011 [accessed 1 February 2024]

33. Welsh Government. Welsh Index of Multiple Deprivation (full Index update with ranks): 2019. https://www.gov.wales/welsh-index-multiple-deprivation-full-index-update-ranks-2019; 2022 [accessed 1 February 2024]

34. Welsh Government. Welsh Index of Multiple Deprivation 2019: analysis relating to areas of deep-rooted deprivation. https://www.gov.wales/welsh-index-multiple-deprivation-2019-analysis-relating-areas-deep-rooted-deprivation; 2022 [accessed 1 February 2024]

35. Fatjo J, Amat M, Mariotti VM, de la Torre JLR, Manteca X. Analysis of 1040 cases of canine aggression in a referral practice in Spain. J Vet Behav. 2007 Sep;2(5):158–65. 10.1016/j.jveb.2007.07.008

36. Barcelos A, Mills DS, Zulch H. Clinical indicators of occult musculoskeletal pain in aggressive dogs. Vet Rec. 2015 May 2;176(18):465–465. 10.1136/vr.102823

37. Wauthier L, SSPCA, Williams J. Using the mini C-BARQ to investigate the effects of puppy farming on dog behaviour. Appl Anim Behav Sci. 2018;206(75–86). 10.1016/j.applanim.2018.05.024

38. Westgarth C, Reevell K, Barclay R. Association between prospective owner viewing of the parents of a puppy and later referral for behavioural problems. Vet Rec. 2012 May 19;170(20):517–517. 10.1136/vr.100138

39. Maher J, Wyatt T. European illegal puppy trade and organised crime. Trends Organ Crime. 2021 Dec 24;24(4):506–25. 10.1007/s12117-021-09429-8

40. Siegel D, van Uhm D. Illegal dogfighting: sport or crime? Trends Organ Crime. 2021 Dec 10;24(4):563–80. 10.1007/s12117-021-09423-0

41. Maher JA, Wyatt T. Rural-urban dynamics in the UK illegal puppy trade: Trafficking and trade in “man’s best friend.” Int J Rural Law Policy. 2019 Aug 6;9(2). 10.5130/ijrlp.2.2019.6266

42. Milroy KE, Whiting M, Abeyesinghe S. Reporting of suspected dog fighting to the police, Royal Society for the Prevention of Cruelty to Animals and equivalents by veterinary professionals in the UK. Vet Rec. 2018 Nov 10;183(18):567–567. 10.1136/vr.104753

43. Maher J, Pierpoint H. Friends, status symbols and weapons: the use of dogs by youth groups and youth gangs. Crime, Law Soc Chang. 2011 Jun 21;55(5):405–20. 10.1007/s10611-011-9294-5

44. NHS England. National Cost Collection for the NHS. https://www.england.nhs.uk/costing-in-the-nhs/national-cost-collection/; 2023 [accessed 1 February 2024]

45. CDC. Cost of Injury Data. https://www.cdc.gov/injury/wisqars/cost/; 2023 [accessed 1 February 2024]

46. Tulloch JSP, Minford S, Vicky P, Rotheram M, Christley RM, Westgarth C. Paediatric Emergency Department dog bite attendance during the COVID-19 pandemic: an audit at a tertiary children’s hospital. BMJ Paediatr Open. 2021 Apr 2;5(1):e001040. 10.1136/bmjpo-2021-001040

47. BMA. COVID-19: Impact of the pandemic on healthcare delivery. https://www.bma.org.uk/advice-and-support/covid-19/what-the-bma-is-doing/covid-19-impact-of-the-pandemic-on-healthcare-delivery#:~:text=Health services across the UK,being missed with growing frequency; 2023 [accessed 1 February 2024]

48. Nath U, Alam B, Das A, Bakhiet A, Pillai A. The Impact of COVID-19 on the Neck of Femur Fracture Service in a Tertiary Care Hospital in the United Kingdom. Cureus. 2023 Oct 18;15(10):e47298. 10.7759/cureus.47298

